# Top-down inhibition of irrelevant information indexed by alpha rhythms is disrupted in migraine

**DOI:** 10.1101/2021.04.29.21256266

**Authors:** Rémy Masson, Hesham A. ElShafei, Geneviève Demarquay, Lesly Fornoni, Yohana Lévêque, Anne Caclin, Aurélie Bidet-Caulet

**Affiliations:** Lyon Neuroscience Research Center (CRNL), INSERM U1028, CNRS UMR 5292, Université Claude Bernard Lyon 1, Université de Lyon, Lyon, France; Neurological Hospital Pierre Wertheimer, Functional Neurology and Epilepsy Department, Hospices Civils de Lyon, Université Claude Bernard Lyon 1, Université de Lyon, Lyon, France

**Keywords:** Migraine, MEG, Alpha activity, Gamma activity, Attention, Distraction, Hypersensitivity

## Abstract

There is growing evidence that migraine is associated with attentional abnormalities, both during and outside migraine attacks, which would impact the cognitive processing of sensory stimulation. However, these attention alterations are poorly characterized and their neurophysiological basis is still unclear. Nineteen migraineurs without aura and nineteen healthy participants were recruited to perform a task which used visually-cued auditory targets and distracting sounds to evaluate conjointly top-down and bottom-up attention mechanisms. Magnetoencephalography (MEG) signals were recorded. We investigated anticipatory alpha activity (power increase and decrease) and distractor-induced gamma activity as markers for top-down (inhibition and facilitation) and bottom-up attention, respectively. Compared to healthy participants, migraineurs presented a significantly less prominent alpha power increase in visual areas in anticipation of the auditory target, indexing a reduced inhibition of task-irrelevant visual areas. However, there was no significant group difference regarding the alpha power decrease in the relevant auditory cortices in anticipation of the target, nor regarding the distractor-induced gamma power increase in the ventral attention network. These results in the alpha band suggest that top-down inhibitory processes in the visual cortices are deficient in migraine but there is no clear evidence supporting a disruption of top-down facilitatory attentional processes. This relative inability to suppress irrelevant sensory information may be underlying the self-reported increased distractibility and contribute to sensory disturbances in migraine.

## 1 Introduction

If headache is undoubtedly its most prominent symptom, migraine is also at its core a sensory processing disorder [21,35]. Migraine attacks are associated to an aversion to external stimulation across all sensory modalities (photophobia, phonophobia, osmophobia, and allodynia) [4,25,47,53,90,97] and this hypersensitivity persists even in the attack-free period, but to a lesser extent [57,90,91]. The most prominent model to explain these sensory disturbances, particularly during the interictal state, has been the migraineurs’ brain habituation deficits to repeated sensory stimulation [17,86]. However, in the last decade, converging pieces of evidence have suggested that dysfunctional attention processing of sensory inputs may also participate in migraineurs’ hypersensitivity, especially during the attack-free period. Migraineurs self-report attention difficulties in everyday life [15,52] and these difficulties correlate with multimodal sensory sensibility between attacks [52]. Migraineurs display increased electrophysiological markers of attention orienting to incoming stimuli [20,61,64,67], which could be related to the dysfunction of the right temporo-parietal junction (rTPJ) [54,55,61,63]. Abnormal attention orienting has been proposed to participate in the excessive sensory discomfort characteristic of other disorders such as autism and attention deficits disorders [13,36,73,74]. Likewise, a deficient attention filter in migraine may lead to an abnormal management of sensory inputs, exposing migraineurs to a state of hyper-responsiveness.

The brain relies on a balance between top-down and bottom-up attentional processes to select relevant information in an environment rich in sources of sensory stimulation. Top-down attention is voluntary and goal-oriented: it promotes the processing of task-relevant information through facilitatory mechanisms and suppresses the processing of irrelevant information through inhibitory mechanisms [11,12,34,44]. Bottom-up attention is involuntary and stimulus-driven: its role is to maintain responsiveness to unexpected but meaningful events through automatic attention shifts that override goal-directed processes [18,29,68]. Alpha rhythms (7–15Hz) are considered to reflect functional inhibition in task-relevant and irrelevant cortical areas [45,49], with increases and decreases in alpha power possibly reflecting facilitatory and inhibitory top-down process, respectively [26,32]. On the other hand, gamma-band activity (>30Hz) would reflect the activation of the cortical areas from which they are generated [7] and has been consistently associated with feedforward pathways directing information from primary sensory areas to associative areas [8,48]. Gamma rhythms are proposed to participate in bottom-up attentional processes [28,32]. Brain rhythms have been investigated in migraine for decades, providing inconsistent results apart from confirming that the visual cortices are hyperexcitable in this population [16], but they have never been studied in the context of the attention function.

In the present article, we analyze oscillatory activities in magnetoencephalography (MEG) signals during an attention task evaluating conjointly top-down and bottom-up attention, in order to better understand the physiological underpinnings of the attention difficulties associated with migraine. We measure alpha and gamma power as markers of top-down and bottom-up attentional processes, respectively. Based on previous work with other brain signals [61,65], we expected alterations for both top-down and bottom-up mechanisms.

## 2 Material and methods

The present study makes use of the same dataset than a previous study [61], in which stimulus-evoked responses (ERP/ERF) were compared between migraine patients and healthy controls. The task, procedure, and preprocessing of MEG data remain identical. Data from 10 control participants are also part of the study presented in ElShafei et al. (2019) [28].

### 2.1 Participants

25 migraine patients (17 female, 8 male) suffering from migraine without aura were included in this study. Inclusion criteria were age between 18 and 60 years and a diagnosis of migraine with a reported migraine frequency between 2 to 5 days per month. Exclusion criteria for patients comprised migraine with aura, chronic migraine, and migraine preventive medication. Every patient was examined by a neurologist (GD, Hospices Civils de Lyon). As we were interested in studying attention during the interictal state, if the patient had a migraine attack during the 72 hours before the testing session, the session was postponed to an ulterior date. If the patient had a migraine attack during the 72 hours after the session, collected data were not used in the analyses, as it is common practice in neuroimaging studies of migraine [21]. Data from 19 patients (13 female, 6 male) were analyzed: data from 5 patients were discarded because a migraine attack happened in the 72 hours following the recording session and data from 1 patient because the patient failed to perform the task correctly. Migraine participants filled the HIT-6 and the MIDAS scales to assess the severity of the disease [50,84].

19 control participants free of migraine and matched to the patients for sex, age, laterality, education level, and musical practice^1^ were included in this study. Exclusion criteria for all subjects included a medical history of psychiatric or neurological disorder except migraine, ongoing background medical treatment other than contraceptive medication, pregnancy, and hearing disability. All subjects gave written informed consent and received a monetary compensation for their participation. Migraine, especially in chronic forms of the disease, is often associated with increased anxiety and depression [22,51,93]. Participants thus filled the Hospital Anxiety and Depression scale [98] in order to verify that anxiety and/or depression were not confounding variables. All demographic statistics can be found in Table 1.

**Table 1:** Demographics and headache profile of the control and migraine groups. Two control participants did not filled the Hospital Anxiety and Depression (HAD) scale. Mean and standard deviation are provided. Group differences are tested using a non-parametric Mann-Whitney U test. NA: not applicable.

|  | **Migraine** | **Control** | **p-value** |
| --- | --- | --- | --- |
| **Sample size** | 19 | 19 | - |
| **Age (years)** | 32.7 (8.7) | 31.2 (7.8) | 0.53 |
| **Sex (number of female participants)** | 13 (68%) | 13 (68%) | - |
| **Education level (years)** | 15.8 (3.1) | 15.8 (2.2) | 0.99 |
| **Musical practice (years)** | 2.8 (3.3) | 2.8 (3.5) | 0.74 |
| **Laterality (number of right-handed)** | 19 | 19 | - |
| **Anxiety score** | 5.7 (3.5) | 4.6 (2.5) | 0.42 |
| **Depression score** | 2.6 (2.6) | 1.8 (2.0) | 0.31 |
| **Migraine duration (years)** | 16.8 (7.4) | NA | - |
| **HIT-6 score** | 64.2 (7.1) | NA | - |
| **MIDAS score** | 12.8 (12.1) | NA | - |

### 2.2 Task and procedure (Figure 1)

Participants performed the Competitive Attention Test, an audio-visual attention task derived from the Posner Task, designed to produce robust measures of both top-down and bottom-up attention. For a detailed description of the protocol used in the present study, please see ElShafei et al. (2019) [28]. For an in-depth discussion of the rationale behind this paradigm and the behavioral responses elicited during the task, please read Bidet-Caulet et al. (2015) and Masson & Bidet-Caulet (2019) [10,60].

**Figure 1:**
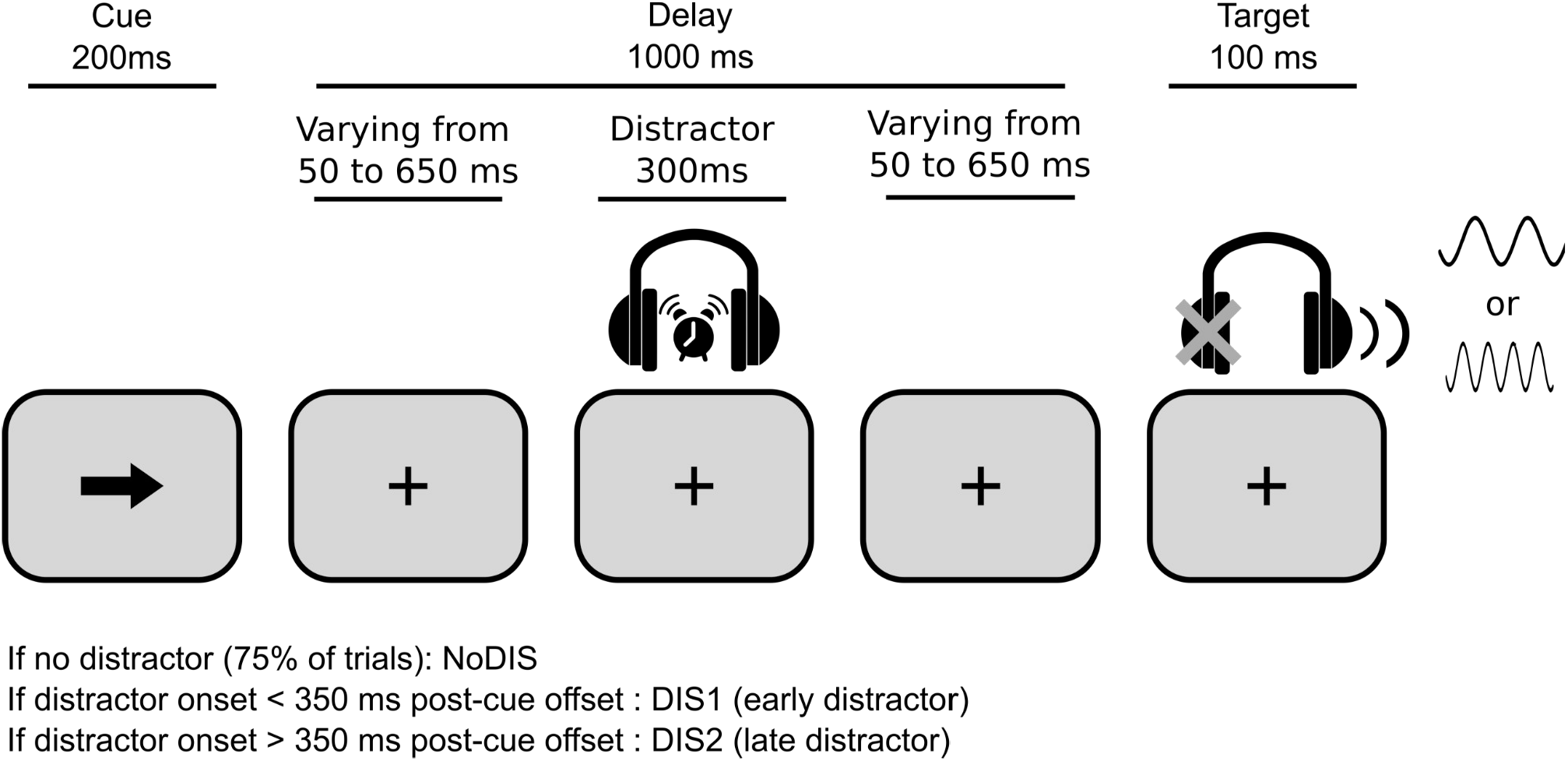
Protocol. The task was to discriminate between a low- and a high-pitched sound, presented monaurally. A visual cue initiated the trial, and was either informative (50%, one-headed arrow) or non-informative (50%, double-headed arrow) about the target ear. 25% of the trials included a distracting sound during the cue-target interval at a random delay after the cue offset: the DIS1 condition corresponds to early distracting sounds (starting 50–350ms after cue offset), the DIS2 condition corresponds to late distracting sounds (starting 350–650ms after cue offset).

In short, all trials comprised a visual cue followed 1000ms later by a monaural auditory target. Half of the time, the cue was a one-headed arrow and was informative on the side of presentation of the target sound; the other half of the time, the cue was a two-headed arrow and was not informative on the side of presentation of the target sound. Participants were asked to discriminate between high-pitched and low-pitched target sounds and to respond as fast as possible using a joystick.

In 25% of the trials, a salient task-irrelevant environmental binaural sound was played at some point between the cue offset and the target onset. If the distracting sound onset was early (50ms to 350ms after cue onset), the trial was categorized as *DIS1*; if the distracting sound onset was late (350ms to 650ms after cue onset), the trial was categorized as *DIS2*. In the 75% remaining trials, no distracting sound was played; trials were categorized as NoDIS.

Full description of the analyses performed on behavioral data can be found in the Supplementary Data.

### 2.3 MEG recording and preprocessing

Simultaneous EEG and MEG data were recorded with a sampling rate of 600Hz during task performance. A 275-channel whole-head axial gradiometer system (CTF-275 by VSM Medtech Inc., Vancouver, Canada) was used to record electromagnetic brain activity (0.016–150Hz filter bandwidth and third-order spatial gradient noise cancellation). Head movements were continuously monitored using 3 coils placed at the nasion and the two preauricular points. EEG was recorded continuously from 7 scalp electrodes and EOG with one bipolar derivation. Please note that EEG data are not presented in this article, as they were only collected for comparison with previous ERP studies, see Masson et al. (2020) [61] for more details.

For each participant, a 3D MRI was obtained using a 3T Siemens Magnetom whole-body scanner (Erlangen, Germany), locations of the nasion and the two preauricular points were marked using fiducials markers. These images were used for reconstruction of individual head shapes to create forward models for the source reconstruction procedures.

MEG data were processed offline using the software package for electrophysiological analysis (ELAN Pack) developed at the Lyon Neuroscience Research Center [1]. Continuous MEG data were bandstop-filtered between 47 and 53Hz, 97 and 103Hz, and 147 and 153Hz (zero-phase shift Butterworth filter, order 3) to remove power-line artifacts. An independent component analysis (ICA) was performed on the 0.1–40Hz band-pass filtered MEG signal to remove eye-movements and heartbeat artifacts. Component topographies and time courses were visually inspected to determine which ones were to be removed through an ICA inverse transformation. 2 to 5 components were removed from the “bandstop-filtered” MEG signal in each participant.

Only trials for which the participant had answered correctly were retained. Trials contaminated with muscular activity or any other remaining artifacts were excluded automatically using a threshold of 2200 femtoTesla for MEG channels. Trials for which the head position differed of more than 10 mm from the median position during the 10 blocks were also excluded from the analyses. For all participants, more than 80 % of trials remained in the analyses after rejection (corresponding to at least 510 remaining trials per subject, including roughly 380 trials with no distractors and 130 trials with distractors).

In anticipation of the baseline correction for distractor-related activity in further analyses, for each distractor onset time range, surrogate distractors were created in the *NoDIS* trials with similar distribution over time than the real distractors.

### 2.4 Time-frequency analyses

All further analyses were conducted using the Fieldtrip MATLAB toolbox (MATLAB 2015A version, Fieldtrip release version 20151231 [72]). This study is focusing on cue-induced alpha and distractor-induced gamma activities. Definition of time-frequency bands and time-windows of interest are based on previous works with a similar paradigm in healthy young adults by Elshafei et al. (2018, 2019) [26,28]. Based on these previous studies, we expected the following pattern of modulations of oscillatory power: (1) decreased power in the low-alpha frequency band (7–11Hz), notably in the motor and auditory cortices, in anticipation of the target sound, reflecting top-down facilitatory processes in the task-relevant areas; (2) increased power in the high-alpha frequency band (11–15Hz) in the occipital cortices in anticipation of the target sound, reflecting top-down inhibitory processes of task-irrelevant visual areas; (3) increased power in the gamma frequency band in the ventral attention network following the presentation of the distracting sound, reflecting the triggering of bottom-up attentional processes.

### a Sensor-level activity

For each single event, the corresponding time-locked response (event-related field) was removed in order to analyze only induced activity free from any evoked activity. Oscillatory power was calculated using Morlet wavelet decomposition with a width of four cycles per wavelet (m = 7; [85]) at center frequencies between 1 and 150Hz, in steps of 1Hz.

For cue-related alpha activity, baseline correction was performed by computing relative change between activity (low-alpha: 7 to 11Hz; high-alpha: 11 to 15Hz, 0 to 1800ms post-cue) and baseline activity (−600 to -200ms pre-cue, averaged over time). For distractor-related gamma-activity, baseline correction was performed by computing relative change between activity (60 to 100Hz, 0 to 350ms post-distractor) in response to distractors vs. surrogate distractors.

Then, for each frequency band, the baseline-corrected activity of interest of the migraine group was contrasted to the control group’s one using a non-parametric cluster-based permutation analysis [58], a statistical strategy that controls for multiple comparisons in time and sensor space dimensions.

### b Source-level activity

First, for each event, the time-locked response was removed in order to analyze only induced activity free from any evoked activity. Then, we utilized the frequency–domain adaptive spatial technique of dynamical imaging of coherent sources (DICS, [38]) in order to reconstruct alpha and gamma activities in the source space dimension. Cross-spectral density (CSD) matrices were calculated using the multitaper method from -600 to 2000ms relative to cue onset (lambda 5%) with a target frequency of 11 (± 4)Hz for NoDis trials, and from -100 to 350ms relative to distractor onset (lambda 5%) with a target frequency of 80 (±20)Hz for all trials. For each participant, an anatomically realistic single-shell headmodel based on the cortical surface was generated from individual head shapes [71]. A grid with 0.5cm resolution was normalized on an MNI template, and then morphed into the brain volume of each participant. Leadfields for all grid points along with the CSD matrix were used to compute a common spatial filter that was used to estimate the spatial distribution of power for the time-frequency window of interest.

In order to estimate source-level activity for each event, we contrasted baseline activity (either pre-stimulus activity for cue-related activities or surrogate distractor-related activity for the real distractor-related activity) to the activity of interest, using non-parametric cluster-based permutations tests which control for multiple comparisons in the source space dimension [58]. The choice of time-frequency windows of interest was informed by the results from previous studies using this paradigm [26–28]. High and low alpha-band activities were investigated between 700 and 1100ms post-cue onset (i.e., in anticipation of the target sound that was played at 1200ms post-cue). Gamma-band activity was investigated between 100 and 300ms after the distracting sound onset.

Then, baseline activity was subtracted from the activity of interest and the resulting difference was contrasted between control and migraine participants using non-parametric cluster-based permutations tests.

## 3 Results

Full description of the behavioral results can be found in Masson et al. (2020) [61] or in the Supplementary Data. In summary, there were no significant group difference neither in general task performance, nor in the behavioral proxies of top-down attention, bottom-up attention and phasic arousal (Supplementary Figure 1).

### 3.1 Cue-elicited alpha-band activities

#### a Anticipation of the target sound: alpha-band activities (Figure 2)

As previously observed in healthy young adults [10,26,27,42,60], the anticipation of the auditory target led, in the control group, to two distinct spatio-temporal patterns for the low-alpha (7–11Hz) and high-alpha frequency (11–15Hz) bands at the sensor level (Figure 2).

**Figure 2:**
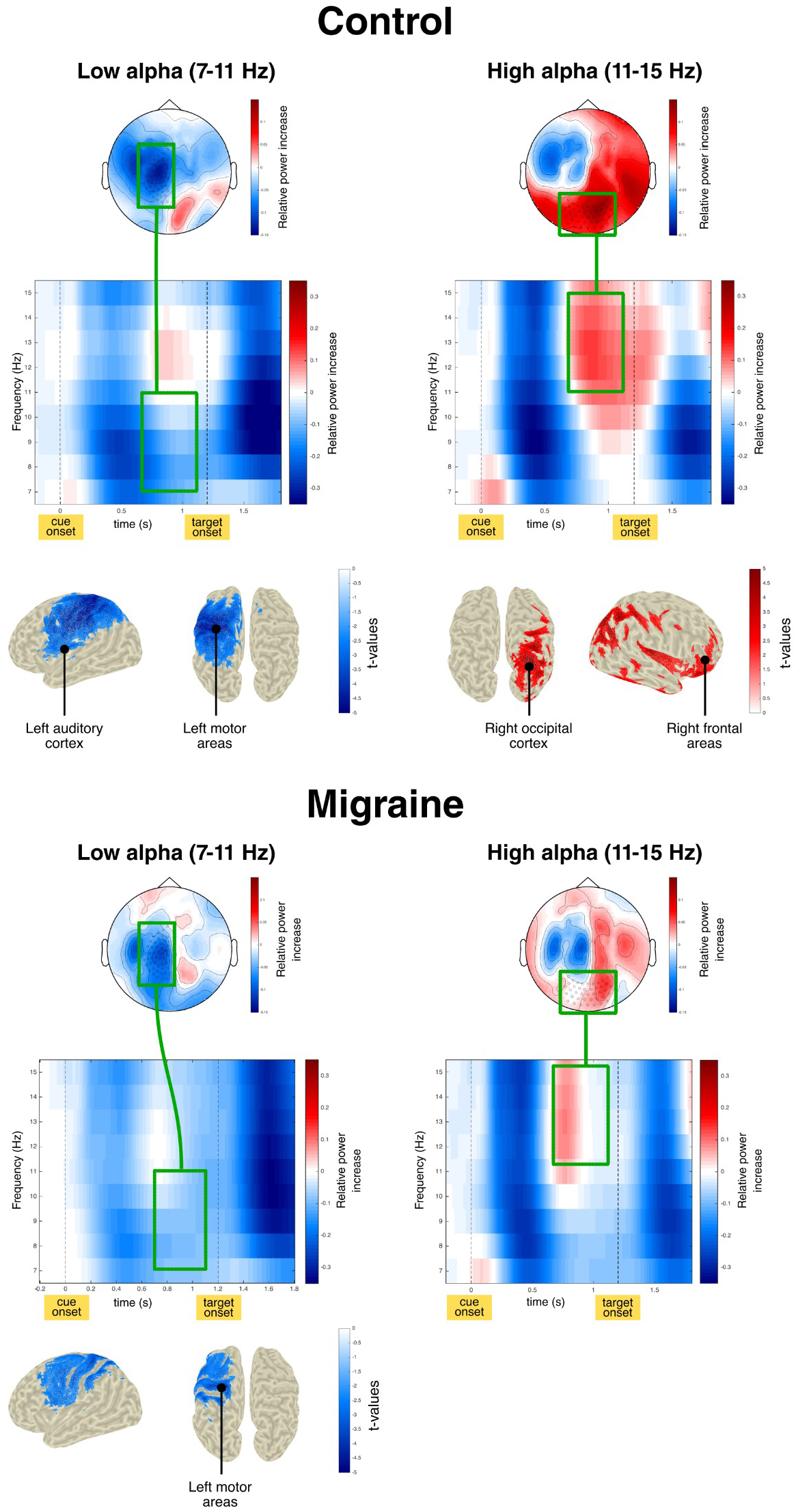
Cue-related low- and high-alpha activity. For each group, the low-alpha (7–11Hz) activity is presented on the left panel, the high-alpha (11–15Hz) is presented on the right panel. (top) Scalp topographies of the baseline-corrected low- or high-alpha power during target anticipation (time-window of interest: 0.7 to 1.1s post-cue onset; baseline window: -0.6 to -0.2 pre-cue onset). (middle) Time-frequency visualization of baseline-corrected alpha power measured at the sensors highlighted by black circles in the topographic maps. (bottom) Distributions of *t* values, masked at corrected *p* < 0.05, from cluster-based permutation tests (one-tailed tests, cluster formation threshold at *p* < 0.05) contrasting low- or high-alpha activity during target anticipation against baseline activity at the source level.

At the sensor level, both the control and migraine groups presented a decrease of low-alpha power over left central and temporal sensors in anticipation of the target sound. Source analysis indicated that this decrease of low-alpha power corresponded to a significant cluster including left motor areas and the left auditory cortex in the control group (p=0.018) and mostly left motor areas in the migraine group (p=0.029).

Control participants also presented an increase of high-alpha power over the occipital and right temporo-parietal sensors starting from 700ms post-cue onset. Source analysis indicated that this increase of high-alpha power corresponded to a significant cluster located only in the right hemisphere and which extended over occipital and parietal areas, the auditory cortex and the orbitofrontal cortex (p=0.007). In the migraine group, the increase of high-alpha power over occipital sensors was much more short-lived. Source analysis did not confirm the presence of a significant pattern of high-alpha power increase (p=0.091).

### b Group differences in alpha-band activities (Figure 3)

At the sensor-level, cluster-based permutation analysis indicated that migraineurs presented less high-alpha (11–15Hz) power in anticipation of the auditory target and during target processing. This corresponded to a significant cluster over occipital sensors, ranging from 900 to 1600ms (p=0.048). Based on this result, high-alpha activity during the 900 to 1600ms time-window was reconstructed, in order to localize the group effect. Source analysis indicated that the larger increase of alpha power observed among control participants emerged from a cluster including bilateral occipital cortices and the right dorso-frontal cortex (p=0.043) (Figure 3).

**Figure 3:**
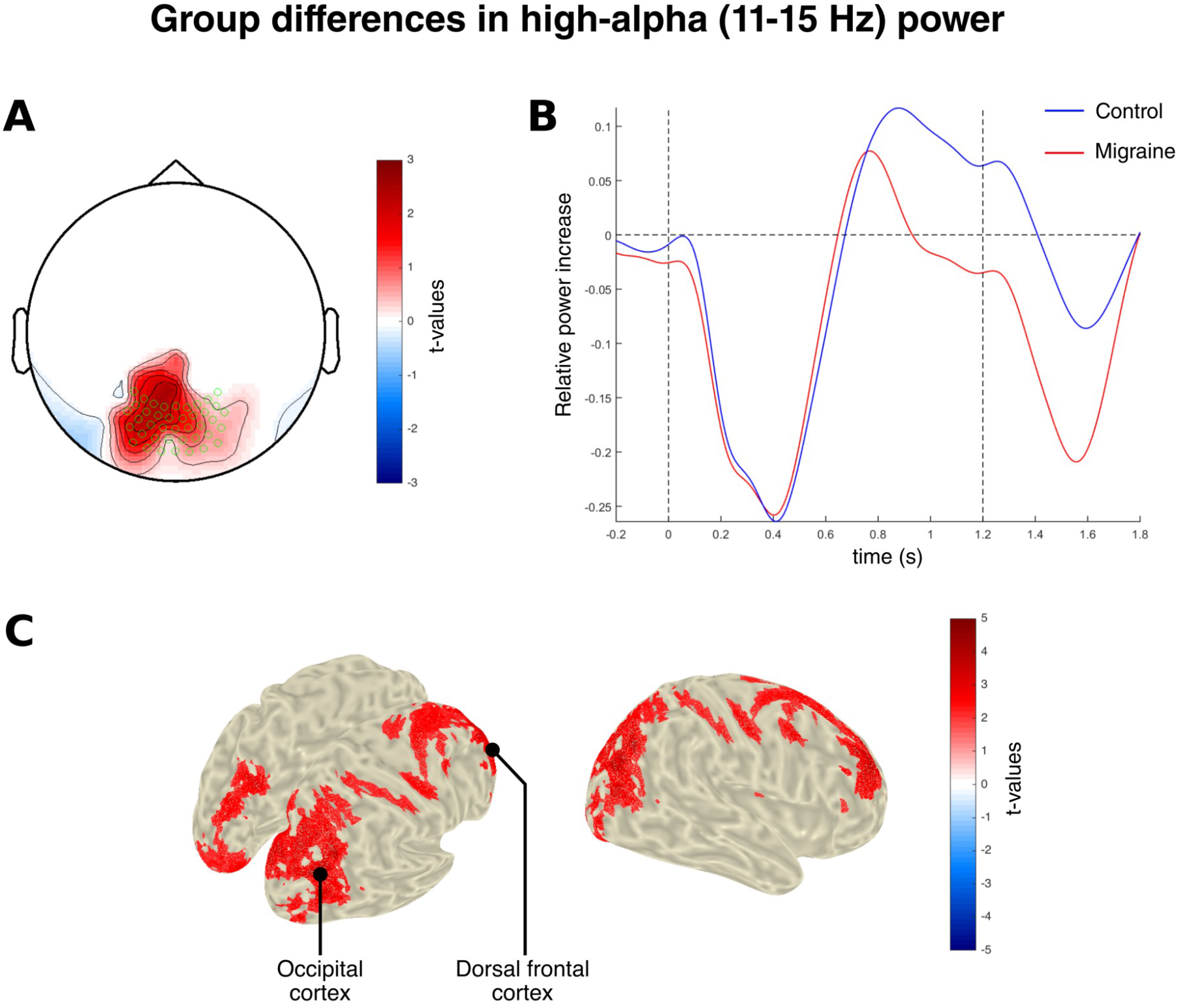
Group differences in cue-related high-alpha activity. (A) Scalp topography of *t* Values from cluster-based permutations contrasting baseline-corrected high-alpha (11–15Hz) activity between the control group and the migraine group at the sensor-level (time-window of interest: 0.9 to 1.6 s post-cue onset; baseline window: -0.6 to -0.2 pre-cue onset). (B) Time-courses of baseline-corrected high-alpha power at occipital sensors (highlighted with green circles in the topographic map) for the control and migraine groups. (C) Distributions of *t* values, masked at corrected *p* < 0.05, from cluster-based permutation tests (two-tailed test, cluster formation threshold at *p* < 0.025) contrasting baseline-corrected high-alpha activity during the time-window of interest between the control group and the migraine group at the source-level (positive values indicate a stronger activity in the migraine group).

No significant group difference was found in the low-alpha band at the sensor-level (p=1). In order to confirm this null result, low-alpha activity in anticipation of the target (700 to 1100ms time-window) was reconstructed. Source analysis showed no significant difference in low-alpha power between the migraine and control groups (p>0.31).

### 3.2 Distractor-elicited gamma-band activity (Figure 4)

As previously observed in healthy young adults [27,28], the distracting sound induced, in both groups, an increase of gamma power (60-100Hz) between 100 and 300ms at the sensor-level. This increase of gamma power was visible over a large number of sensors but was maximal over a left and a right focal clusters of temporo-parietal sensors. In the control group, source analysis indicated that this increase of gamma-band power corresponded to a significant cluster including the bilateral temporo-parietal junctions, both auditory cortices and the right dorso-lateral prefrontal cortex (p<0.001). In the migraine group, source analysis indicated that the increase of gamma-band power corresponded to four significant clusters including the right temporo-parietal junction, the right sensorimotor cortex and both auditory cortices (p<0.01 for all four clusters).

**Figure 4:**
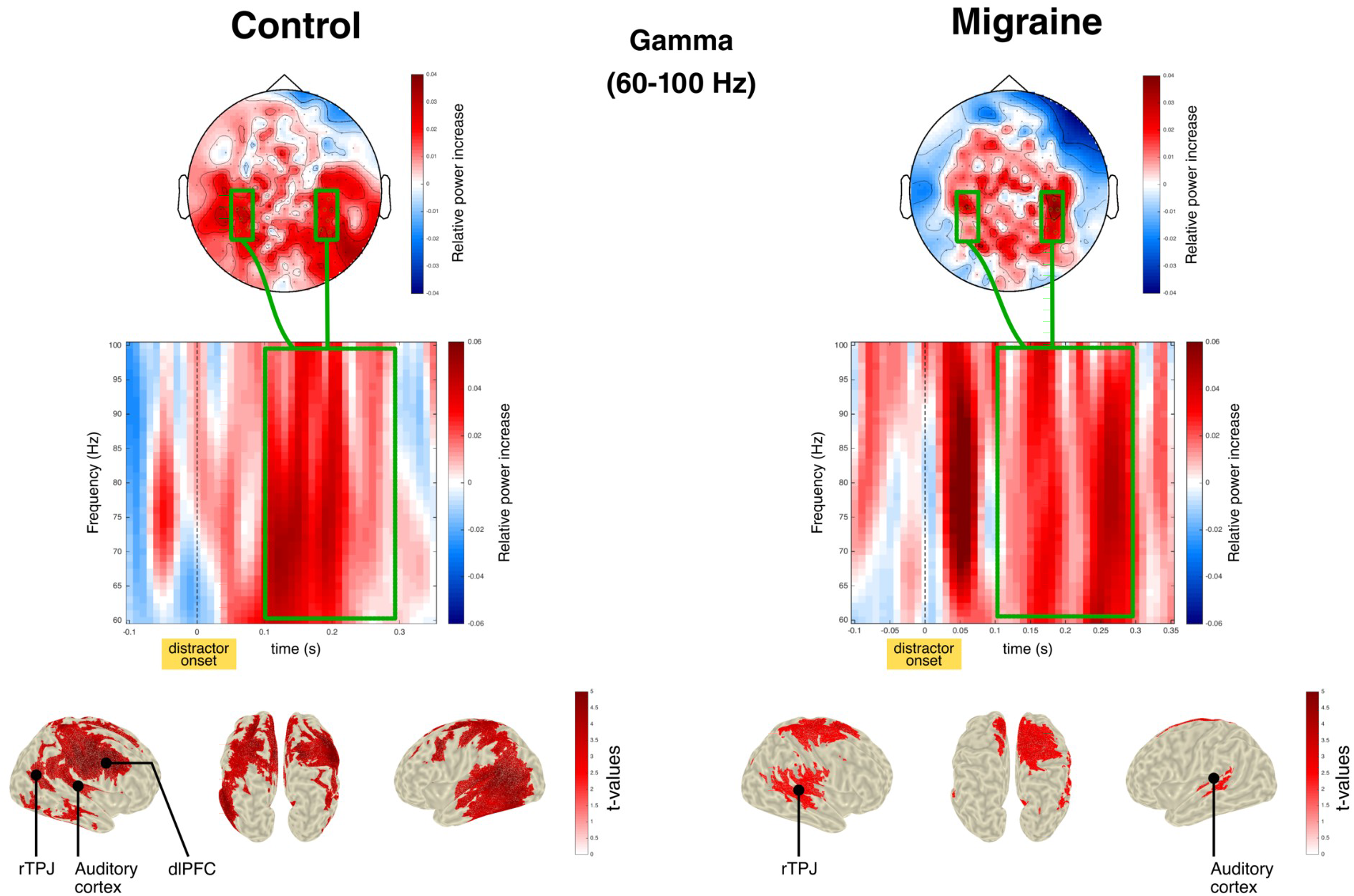
Distractor-related gamma activity. (top) Scalp topographies of the baseline-corrected gamma power (frequency of interest: 60-100Hz; time-window of interest: 0.1 to 0.3s post-distractor onset). (middle) Time-frequency visualization of baseline-corrected gamma power measured at the temporo-parietal sensors highlighted with green circles in the topographic map. (bottom) Distributions of *t* values, masked at corrected *p* < 0.05, from cluster-based permutation tests (one-tailed tests, cluster formation threshold at *p* < 0.005 for the control group, *p* < 0.01 for the migraine group) contrasting contrasting real and surrogate distractor gamma activity during the time-window of interest at the source level.

No significant group difference in gamma power between 100 and 300ms was found at the sensor, not at the source level (p=1).

## 4 Discussion

Migraine has been associated with attentional abnormalities through subjective reports [15,52,78], neuropsychological tests [89,92], or EEG and event-related potentials (ERPs) studies [65]. However, little is known on the functional role of brain oscillations in the disruption of attention in migraine. In the present study, we investigated alpha activity during top-down anticipatory processes and gamma activity during bottom-up processing of distracting auditory stimuli, in migraine patients without aura. Migraineurs displayed dysfunctional top-down inhibitory processes, as revealed by a reduced alpha synchronization (power increase) in visual areas in anticipation of an auditory target, in comparison to control participants. In contrast, top-down facilitatory processes and bottom-up processes were found unaffected in the migraine group.

### 4.1 Top-down anticipatory processes in migraine

There is growing consensus that alpha rhythms reflect active functional inhibitory processes through the modulation of neuronal excitability [45,49]. During attention tasks, attending to the location, feature or timing of a stimulus results in a decrease of alpha power in sensory areas relevant for its processing, and in an increase of alpha power in irrelevant sensory areas. These alpha modulations have been described in both the visual [46,59,75–77,82] and the auditory [69,95,96] modalities. During multisensory paradigms such as the one used here, alpha power has been reported to increase in brain areas processing the unattended sensory modality and to decrease in relevant sensory areas [23,31,33,62]. It has been specifically observed in a previous study in young healthy adults that the anticipation of an auditory target is associated to an increase of high-alpha (11–15Hz) power in the task-irrelevant occipital cortex concomitant to a decrease of low-alpha (7–11Hz) power in the task-relevant auditory cortex, reflecting distinct inhibitory and facilitatory attentional mechanisms [26]. Inhibition of the visual pathway was found functionally relevant as behavioral performance positively correlated with the increase of alpha power in the occipital cortex [26].

In the present study, using a similar paradigm, we confirm in the control group that the anticipation of an auditory target leads to: (1) a decrease of low-alpha power in the auditory cortex, interpreted as a facilitation of auditory processing; and (2) an increase of high-alpha power in the occipital cortex, interpreted as an inhibition of visual processing. Compared to healthy control participants, migraineurs present a less pronounced and more short-lived alpha power increase in the occipital cortex in anticipation of the auditory target and during target processing. Migraineurs appear to be less efficient at suppressing the task-irrelevant visual pathways, which suggests that migraine is associated with deficient top-down inhibitory processes. Abnormal excitability of occipital cortices in migraine has already been proposed to be a feature of the migraine brain [6]. Notably, unlike healthy participants, migraineurs present highly synchronized steady-state activity in the alpha band during flickering stimulation [3,87,88], suggesting difficulties to suppress repetitive visual stimulation.

Migraineurs also present a decreased alpha power in dorsal prefrontal areas compared to control participants. The dorsal prefrontal cortex has been consistently associated with goal-oriented, top-down attentional processes [5,19], and as part of the dorsal attention network it has been proposed that it coordinates alpha power modulation in sensory cortices [14,24,43,56,69,96].

By contrast, no significant difference was observed between the two groups in terms of low-alpha decrease in the auditory cortex. One could have expected an inadequate alpha decrease in the auditory cortex in anticipation of an auditory target. This result suggests that top-down facilitatory processes might be preserved in migraine, as task-relevant sensory pathways appear to be functionally unaffected here.

In previous works investigating top-down attention in migraine, migraineurs displayed different patterns regarding facilitatory and inhibitory attentional mechanisms. [66] On the one hand, migraineurs appear to present a better-than-normal ability to facilitate in a top-down fashion the processing of visual [66] or auditory target stimuli [61]. Group differences were only observable during target processing, not during the anticipatory period, suggesting that it is mainly “reactive” facilitatory processes that are exacerbated, not “pro-active” anticipatory processes. On the other hand, migraine appears to be associated with an impairment of visual noise suppression [94], and with a decreased ability to suppress unattended inputs in the periphery [63,66]. In conclusion, migraine seems to be characterized by a normal or even increased propensity to facilitate the processing of relevant inputs; but importantly also by a deficient ability to pro-actively suppress irrelevant information. However, based on these results and the previous literature, it remains unclear if the dysfunction of top-down inhibition in migraine is specific of the visual cortices, or if it extends to other sensory regions.

### 4.2 Bottom-up attentional processes in migraine

Gamma activity has been associated with attention mechanisms. During attention tasks, gamma activity behaves opposite to alpha rhythms, as gamma power is enhanced in task-relevant areas [37]. Gamma activity is proposed to reflect feed-forward, bottom-up processes, contrary to alpha rhythms which are more closely associated with feedback, top-down mechanisms [32,80].

As expected [28], we observe that distracting sounds trigger an increase of gamma power within the ventral attention network (VAN) and auditory cortices in healthy and migraine participants. This distractor-induced gamma burst is interpreted as being the physiological manifestation of attention capture by distracting sounds, as the VAN is considered to underlie stimulus-driven, bottom-up attention mechanisms [18,19,79]. In the present study, contrary to our a priori hypotheses, migraine patients do not appear to present an enhanced recruitment of the VAN in response to a salient, unexpected sound, as assessed with the study of gamma-band activity.

In our previous analysis of the same data set [61], we observed that migraineurs displayed increased event-related responses to distracting sounds, namely the orienting response of the N1 and the re-orienting negativity (RON), and presented during the RON an increased recruitment of the right temporo-parietal junction (rTPJ). The rTPJ is a crucial node of the VAN linked to bottom-up attention orienting, and is also strongly involved in attention shifting [18]. The absence of group difference in the distractor-elicited gamma activity in the VAN might seem surprising in light of previous results. However, distraction is not limited to the sole attention capture, it is generally understood as a three-stage process [30,40]. First, a change detection mechanism reflected by the orienting component of the N1 is elicited by sudden, infrequent sounds [2,9,70]; this detection mechanism would trigger an attention shift towards the unexpected sound, through the activation of the VAN [28,83] and reflected by the P3a [29]; finally, the participants would reorient their attention back to the task at hand as reflected by the RON [81]. These three steps are not always strongly coupled and their respective markers may vary independently [39,40]. In migraine, the second stage – attention capture – might not be negatively impacted, as evidenced by the typical distractor-elicited P3a [61] and gamma activity in the VAN. However, the change detection mechanism could be more easily triggered in migraine, as evidenced by the heightened N1, without necessarily leading to an exacerbated orienting response. The heightened RON and the increased recruitment of the rTPJ following distractors could then be seen as a compensatory top-down mechanism for shifting attention back to the task, which allows migraineurs to better recover from distraction. A typical bottom-up attention capture despite a more prominent change detection mechanism would explain why migraineurs self-report being more easily bothered by salient sounds [52], but do not display significantly worse behavioral costs by distractors during the present task.

### 4.3 Conclusions & Clinical Perspectives

In a world filled with multiple sources of sensory stimulation, top-down selective attention enables us to focus on relevant aspects of the environment and ignoring those which are irrelevant, while bottom-up attention allows to remain aware of unattended but potentially meaningful events. Based on the present results, migraine seems to be associated with a deficient recruitment of top-down inhibitory mechanisms as migraineurs appear to be less capable of inhibiting visual areas when they are not relevant to the task. However, there is no evidence in the present data that top-down facilitatory mechanisms are negatively impacted in migraine, in agreement with the previous literature on the subject. The recruitment of the ventral attention network has not been found to be abnormal, perhaps indicating that migraineurs somehow manage to maintain normal bottom-up attention capture to unexpected sounds despite a general state of hyperresponsiveness.

The apparent inability of migraineurs to suppress irrelevant information may be part of the neurophysiological underpinning of their complaints of attention difficulties and increased distractibility in their daily life [15,52,65,78]. The characterization of attentional alterations associated with migraine might inform therapeutical strategies to improve their daily life and possibly reducing the attack frequency. Based on the present results, migraineurs would not have major problems to focus on their work but would fail to effectively suppress irrelevant sensory inputs. Clinical training specifically focusing on distraction and noise suppression may be found useful for symptoms management. Finally, deficient top-down inhibitory mechanisms may play a role in the multimodal hypersensitivity during both the headache phase and the pain-free period [52,57,90,91]. Further research is needed to establish a clearer relationship between sensory complaints and the disruption of the attention processing of sensory stimuli in migraine.

## Data Availability

The data that support the findings of this study are not publically available as participants did not consent to data sharing

## 5 Supplementary Material

### 5.1 Analysis of behavioral data

The Competitive Attention Test (CAT) has been shown to provide robust behavioral indexes of top-down attention, bottom-up attention capture, and phasic arousal in various populations: healthy young adults [10,26,28,60], the elderly [27] or healthy children [41]. In these previous studies, participants were faster in trials with informative cues compared to those with uninformative cues: the difference in reaction time (RT) between *informative* and *uninformative* trials was considered a proxy for top-down anticipatory attention. Participants were faster in trials with an early distractor than in those without distracting sound: the difference in RT between NoDIS and DIS1 trials was considered a proxy for an increase of phasic arousal. Finally, participants were slower in trials with a late distractor compared to those with an early distracting sound: the difference in RT between DIS2 and DIS1 trials was considered a proxy for bottom-up attention capture.

Analyses of behavioral data were already performed in a previous article using this dataset [61]. Only trials with a correct response were retained and median RT for each subject and condition were computed. A repeated-measures ANOVA with CUE category (2 levels: *uninformative, informative*) and DISTRACTOR condition (3 levels: *NoDIS, DIS1, DIS2*) as within-subject factors and GROUP category (2 levels: *control, migraine*) as a between-subject factor was conducted. To correct for possible violations of the sphericity assumption, Greenhouse-Geisser correction was applied to resulting p-values. Post-hoc comparisons were conducted using t-tests followed by a Bonferroni correction. Statistical analyses were conducted using the software JASP (version 0.9).

### 5.2 Behavioral results (Supplementary Figure 1)

Main effects of the DISTRACTOR and CUE categories were found significant. Participants responded significantly faster in the *informative* condition than in the *uninformative* condition. Participants responded significantly faster in trials with an early distracting sound *(DIS1)* compared to trials without distracting sound *(NoDIS)*. Participants responded significantly slower in trials with a late distracting sound *(DIS2)* compared to trials with an early distracting sound *(DIS1)* or even compared to trials without distracting sound *(NoDIS)*. Neither the main effect of the GROUP category nor the interactions of GROUP with CUE or DISTRACTOR category were found significant. Full description of the behavioral effects can be found in Masson et al. (2020) [61].

## Tables & Figures

**Supplementary Figure 1:**
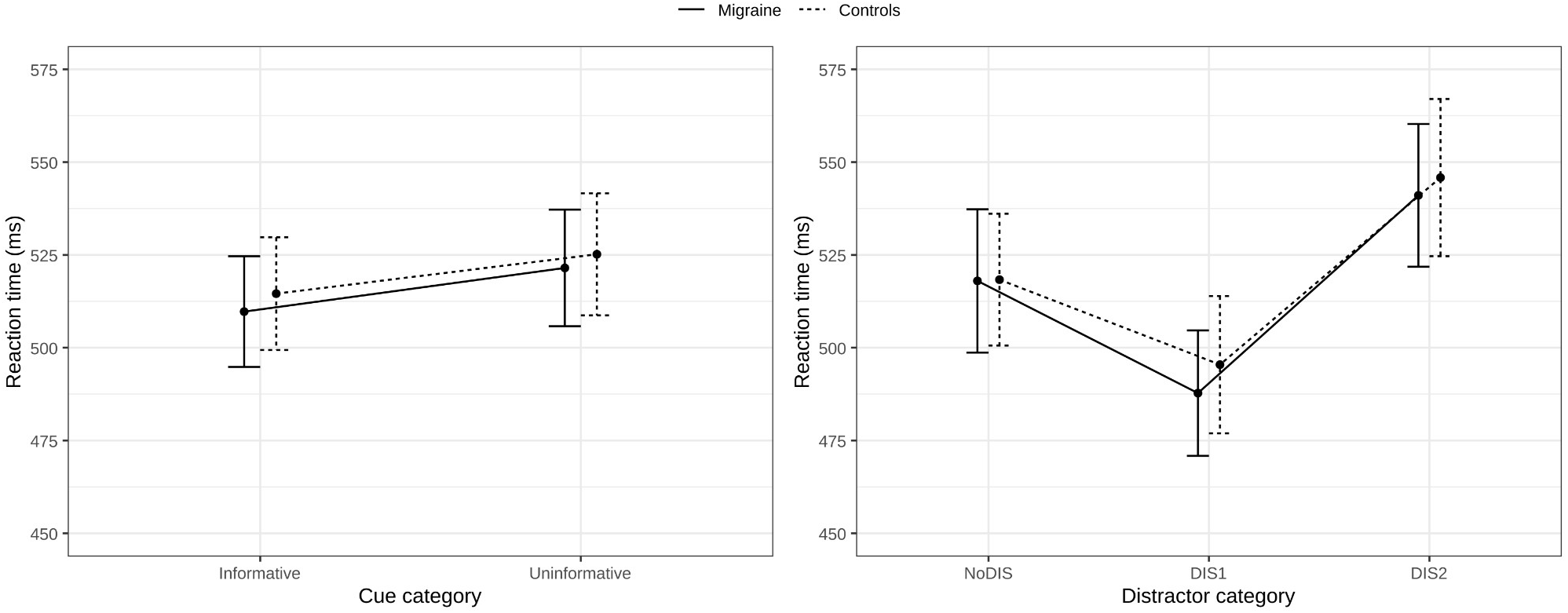
Behavioral results. Reaction time (RT) as a function of the CUE and GROUP category on the left panel, RT as a function of the DISTRACTOR and GROUP category on the right panel. Participants present a significant effect of the cue informational value (*uninformative* > *informative*), a significant effect of phasic arousal (*NoDIS* > *DIS1*) and a significant effect of attention capture (*DIS2* > *DIS1*). A repeated-measures ANOVA showed no significant interaction of the GROUP with the CUE or with the DISTRACTOR category. See Masson et al. (2020) for more details [61].

## Footnotes

1 Pitch discrimination is required in the task described below, and is an ability increasing with musical practice.

